# STAT4 Phosphorylation of T-helper Cells predicts surgical outcomes in Refractory Chronic Rhinosinusitis

**DOI:** 10.1101/2023.12.11.23299743

**Authors:** Michael R. Abidin, Oral Alpan, Matthew Plassmeyer, Lina Kozhaya, Denise Loizou, Mikail Dogan, Zachary Upchurch, Nathan P Manes, Aleksandra Nita-Lazar, Derya Unutmaz, Søren Ulrik Sønder

## Abstract

Objective: Chronic rhinosinusitis (CRS) impacts an estimated 5% to 15% of people worldwide, incurring significant economic healthcare burden. There is a urgent need for the discovery of predictive biomarkers to improve treatment strategies and outcomes for CRS patients.

Study design: Cohort study of CRS patients and healthy controls using blood samples. Setting: Out-patient clinics.

Methods: Whole blood samples were collected for flow cytometric analysis. Mechanistic studies involved the transfection of human primary T cells and Jurkat cells.

Results: Our analysis began with a 63-69 year-old female patient diagnosed with refractory CRS,. Despite undergoing multiple surgeries, she continually faced sinus infections. Whole exome sequencing pinpointed a heterozygous IL-12Rb1 mutation situated in the linker region adjacent to the cytokine binding domain. When subjected to IL-12 stimulation, the patient’s CD4 T-cells exhibited diminished STAT4 phosphorylation. However, computer modeling or T-cell lines harboring the same IL-12 receptor mutation did not corroborate the hypothesis that IL-12Rb could be responsible for the reduced phosphorylation of STAT4 by IL-12 stimulation.

Upon expanding our investigation to a broader CRS patient group using the pSTAT4 assay, we discerned a subset of refractory CRS patients with abnormally low STAT4 phosphorylation.

The deficiency showed improvement both in-vitro and in-vivo after exposure to *Latilactobacillus sakei* (aka *Lactobacillus sakei*), an effect at least partially dependent on IL-12.

Conclusion: In refractory CRS patients, an identified STAT4 defect correlates with poor clinical outcomes after sinus surgery, which can be therapeutically targeted by *Latilactobacillus sakei* treatment. Prospective double-blind placebo-controlled trials are needed to validate our findings.

## Introduction

The intricate web of cellular interactions and cytokine signaling pathways plays a pivotal role in orchestrating the immune response to pathogens. Central to this network is the IL-12/IFN-γ axis, a well-recognized T-helper 1 (Th1) pathway in mediating protective immunity against intracellular pathogens [1]. Interleukin-12 (IL-12), produced predominantly by antigen-presenting cells like dendritic cells and macrophages [2], induces the production of interferon-gamma (IFN-γ) from natural killer cells and T lymphocytes [3,4]. IFN-γ, in turn, acts to stimulate the innate and adaptive arms of the immune system, promoting cellular immunity and controlling pathogen proliferation. IL12Rβ1 is a subunit of both the IL-12 and IL-23 receptor complex and binds to the p40 subunit of IL-12 and IL-23. Upon infection with intracellular bacteria, phagocytes are activated and secrete cytokines including IL-12, a signature cytokine for Th1 responses. Following binding of IL-12 to its receptor, STAT4 is phosphorylated which is essential for IFNγ production[1,5,6].

Disruption of this axis could compromise the ability of the immune system to handle microbial challenges, leading to persistent infections and prolonged inflammation. Indeed, a balanced IL-12/IFN-γ response is essential to clear pathogens, maintain mucosal homeostasis, and prevent tissue damage in the sinuses[1]. Furthermore, recurrent infections, which denote a failure of the immune system to provide lasting protection after initial exposure to a pathogen, may also be linked to perturbations in the IL-12/IFN-γ pathway. An impaired or inadequate IL-12-driven IFN-γ response may not only decrease resistance to primary infections but might also reduce immune memory, rendering individuals more susceptible to recurrent bouts of infections despite adequate surgical intervention [1,7–13]. Although T-cell defects are the most common immune disorder in chronic rhinosinusitis (CRS), clinical workup towards its identification is rarely performed.

Various dietary supplements have been demonstrated to enhance Th1 responses. Among them included are various lactobacillus probiotic supplements. Although the role of lactobacillus containing supplements, taken orally or topically, is unproven in patients with CRS, small sized studies and anecdotal reports did find correlations with clinical outcomes in the surgical healing process [14–20]. Data from preliminary studies have shown mixed results in patients receiving probiotics containing lactobacilli species. Lactobacilli have the potential of inducing IL-12 from macrophages [21–23].

Our investigation began with a case of refractory chronic rhinosinusitis (RCRS) associated with a heterozygous IL-12 receptor mutation. Despite observing decreased STAT4 phosphorylation in response to IL-12 stimulation in both the patient and her children, the mutation itself was not proven to be the cause. Further research revealed this decreased phosphorylation to be a common characteristic in a broader RCRS patient cohort. Insights into the IL-12/IFN-γ signaling pathway’s role in CRS may unveil new therapeutic targets to enhance immunity, alleviate chronic inflammation, and decrease recurrent infections.

## Materials and Methods

### Patients and ethical statement

The studies involving human participants were reviewed and approved by Western Institutional Review Board (WIRB) Protocol #1285028. Written, informed consent was obtained from the individuals for the publication of any potentially identifiable images or data included in this article. Individuals undergoing steroid or MoAB treatments targeting the IL-12 pathway were excluded from the study. The grouping of CRS and RCRS patient was done according to published guidelines[24,25].

Lactobacilli supplements and administration: *Lactobasillus sakei* proBio65 as a nasal rinse 1/4^th^ teaspoon of Lanto Health powder (240 mg) in 2 tablespoons of normal saline nasal rinses once a day and 240 mg twice a day in food.

Genetics: Whole Exome Sequencing was carried out by GeneDX (Gaithersburg, MD). Subsequently Sanger sequencing was carried out by Macrogen (Rockville, MD).

### Flow cytometry and cell culture

Staining for IL-12Rβ1 expression was performed using antibodies against IL-12Rβ1, CD4, CD20 and CD3. For the Th1 & Th17 assay PBMC were isolated by gradient centrifugation. The percentages of CD4 T-cells capable of producing IFNγ (Th1 cells) and IL-17 (Th17 cells) were measured by stimulating the PBMC with PMA (50ng/ml) and ionomycin (1μg/ml) in the presence of brefeldin A for 4 hours. The cells were then permeabilized and stained with antibodies against CD3, CD4, CD8, CD45RA, CCR6, IFNγ-APC and IL17-PE (BD Bioscience, San Jose, CA). Acquisition was set to 30000 CD3^+^ events. The Th1 mediated IFNγ response was studied by incubating the PBMC for 72 hours in the presents of IL-2 (100 U/ml), IL-12 (100 ng/ml), IL-18 (50 ng/ml), and Dynabeads Human T-Activator CD3/CD28 (10μl/ml). The cells were then fixed, permeabilized and stained with antibodies against CD3, CD45RO, CD4, IFNγ and fixable viability dye 780. Acquisition was set to 20000 CD4^+^ events. The CAP and CLIA validated pSTAT4 stimulation assays were carried out using whole blood stimulated with IL-12 (100ng/ml) for various timepoints, followed by staining with antibodies against CD3, CD4, CD45RO, and pSTAT4(pY693) (BD Bioscience, San Jose, CA). Acquisition was set to 20000 CD4^+^ events with a LLOQ of 2500[26]. All samples were acquired on a 3 laser/10 color BD FACSCanto. The instrument has been CAP (College of American Pathologists) and CLIA validated for clinical diagnostic studies. The individual antibody concentrations were optimized for maximum separation of the populations[27].

One capsule of Super 8 HI-Potency Probiotic from Flora containing *L. acidophilus, L. rhamnosus, L. salivarius, L. plantarum, L. casei, B. bifidum and B. longum* was resuspended in PBS and a total of 10^6^ cells were added to 10^6^ PBMC in the presence or absence of IL-12 neutralizing antibodies (BioRad) and incubated for 18 hours at 37°C. Control samples were stimulated with IL-12 for 2 hours. The experiment was done six times and representative plots are shown. The percentage reduction in pSTAT4 phosphorylation was calculated using the CD45RO^+^/pSTAT4^+^ value as ((IL12AB+lactobacilli)-(unstimulated))*100/((lactobacilli)-(unstimulated))

### Data analysis and modelling

Data analysis of flow cytometry data was performed using FCS Express software (De Novo software, Glendale, CA). All data comparisons were analyzed as paired, two tailed, two-sample unequal variance using Student’s t-test to determine significance. A p-value less than 0.05 was considered significant, * p<0.05,** p<0.01.

Protein structure modeling was performed using AlphaFold [28]. The resulting models were aligned and visualized using Chimera[29].

### Plasmid constructs

Human codon-optimized wild-type IL-12Rb1 and IL-12Rb2 protein sequences were synthesized by MolecularCloud (HG11674-UT and HG10145-UT, respectively) and then cloned into lentiviral plasmid constructs with RFP and GFP reporters, respectively. The overlap extension PCR method was used to generate the mutant IL12Rb1 Q238E protein sequence and clone it into the lentivector. IL-12Rb1 Q238E sequence was then confirmed using sequencing by Eton Bio. Primer sequences are available upon request.

### Lentivirus production

The lentiviruses pseudotyped with vesicular stomatitis virus G protein envelope were generated with HEK293T cells using Lipofectamine 3000 (Invitrogen) according to the manufacturer’s protocol as previously described [30].

### Engineering primary human T cells and Jurkat cells

Healthy adult blood was obtained from AllCells. Primary CD4 T cells were isolated, activated, transduced to overexpress IL12Rb1 wild-type or mutant, and proliferated as previously described [31]. Total PBMC were activated with antiCD3/anti-CD28 dynabeads (Invitrogen) and expanded in IL-2 (10ng/ml) containing media. To generate IL12Rb1 wild-type or mutant and IL12Rb2 overexpressing Jurkat cells, wild-type Jurkats were transduced with the IL-12Rb2 lentiviruses, co-expressing GFP marker, and superinfected with IL-12Rb1 wild-type or IL-12Rb1 Q238E lentiviruses, co-expressing RFP marker, at MOI=3. The infection levels were determined by GFP or RFP expression using flow cytometry analysis. Engineered Jurkat cells were later single-cell cloned with FACSAria Fusion (BD Biosciences). To further confirm IL-12 receptors overexpression, transduced cells were stained with IL-12Rb1 (R&D) and IL-12Rb2 (Biolegend) antibodies. For phospho-Stat4 staining, total resting or activated PBMCs, engineered primary CD4 T or Jurkat cells were stimulated with different concentrations of IL-12 (R&D) for 90 minutes. Cells were collected, stained with fixable viability dye (eBiosciences) and surface markers including IL-12Rb1 (R&D biosystems), CD3, CD4 and CD8 (Biolegend) to identify PBMC populations, washed then fixed with Cytofix buffer (BD) prewarmed to 37°C for 10 min. After spinning down, the cells were permeabilized with ice-cold Phosflow Perm buffer III (BD) on ice for 30 min and were stained with phospho-Stat4 (pY693) antibody (BD) or total Stat4 primary antibody followed by anti-rabbit secondary antibody (both from Invitrogen). Stained cells were analyzed on BD Symphony A5 flow cytometer (BD Biosciences). Flow cytometry data was analyzed using FlowJo software.

### Statistical Analyses and Reproducibility

All statistical analyses were performed, and graphs were prepared using GraphPad Prism V9 software. Each experiment was performed at least three times and non-parametric t-tests were used for statistical significance analyses.

## Results

### Identification of a Th1 defect in a family of patients with RCRS

The patient is a 63-69 year-old Caucasian female with a long-standing history of chronic pansinusitis without polyps, and osteomyelitis of the right maxillary sinus wall. Sinus cultures grew methicillin resistant S. aureus and Klebsiella species on multiple occasions. Patients underwent multiple sinus surgeries and procedures involving ethmoid, maxillary and frontal sinuses. Sinus surgical pathology results show predominantly neutrophilic infiltrates (Figure 1A). The patient’s immune evaluation showed slight hypogammaglobulinemia (400-500 mg/dL range) on multiple occasions along with poor responses to S. pneumoniae serotypes (3/14 of the serotypes with protective levels above 1.3 mcg/ml despite multiple boosters with Pneumovax23). Serum IgM, IgA and IgE levels were within normal limits. The patient’s B cells expressed normal CD27, IgG, IgA, IgM and IgD, arguing against a primary antibody deficiency disorder. T-cells, including T-cell subsets and NK cells were normal for absolute numbers and percentages. Given the poor response to antibiotic therapies, and persistently borderline low IgG, the patient was started on IgG replacement therapy which resulted in partial clinical improvement of the sinus disease.

**Figure 1.**
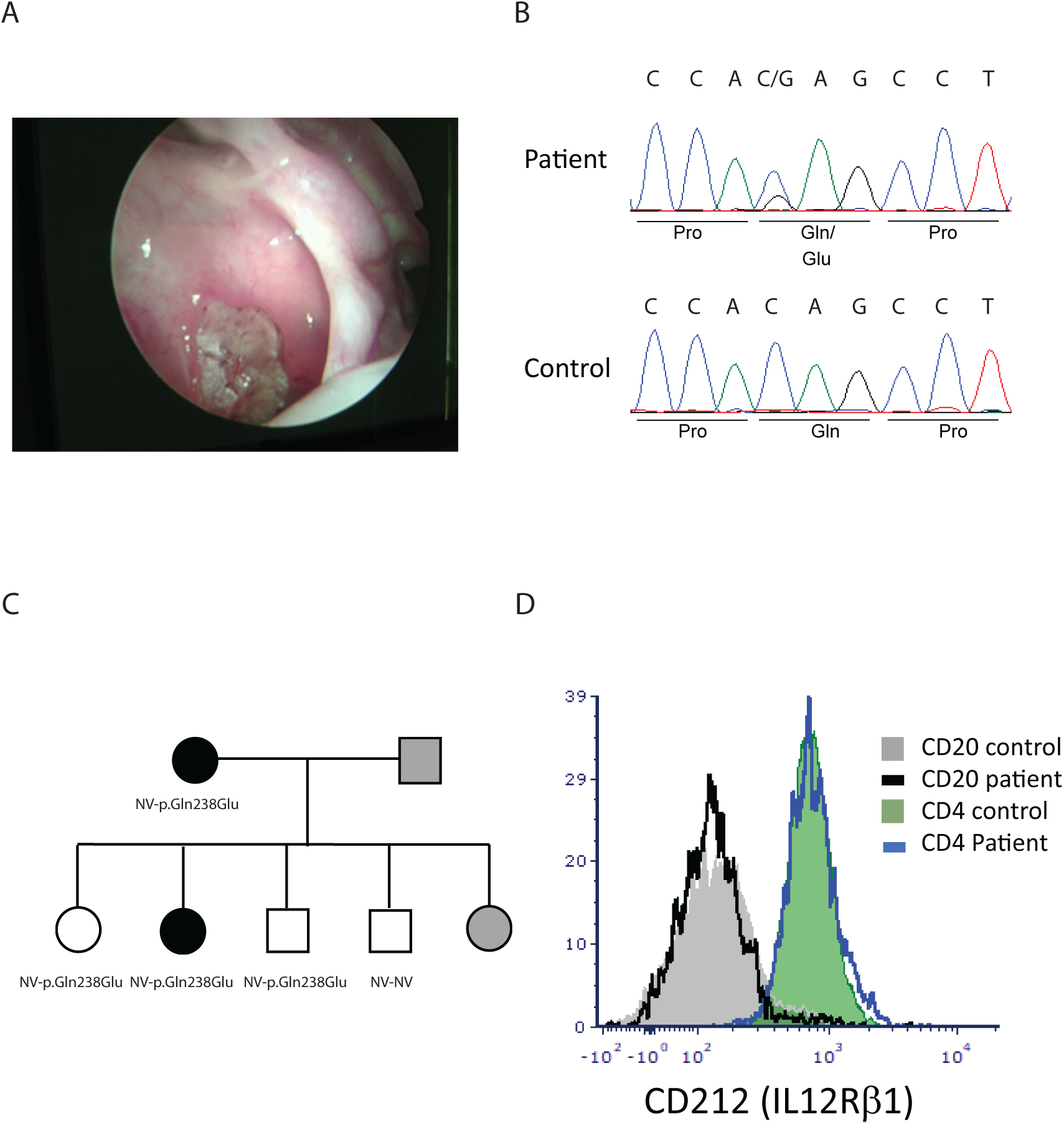
Characterizing of the patient. A) Endoscopic photograph of the patient’s sinus. B) Sanger sequencing of the *IL12Rb1* gene. C) Pedigree of the patient and her family. D) CD212 (IL12Rβ1) expression on CD4 and CD20 lymphocytes.

Since the patient’s clinical presentation was not fully explained by the low serum IgG and only a partial clinical response to the IgG replacement therapy, whole exome sequencing was performed, which revealed a heterozygous mutation in the *IL12RB1* gene, 712 C>G resulting in the p.Gln238Glu variant. The variance was confirmed by Sanger sequencing in the patient as well as in four of her five children (Figure 1B+1C). The p.Gln238Glu variant is classified as a variant of uncertain significance (VUS) in ClinVar[32]. The structure modeling of the molecular defect maps it to the linker region to the cytokine binding domain (Supplemental figure 1). The mutation did not affect the expression level of IL12Rb1 receptor on the cell surface or the ability of CD4 cells to produce IL-17 or IFNγ in response to general activation by PMA plus ionomycin (Figure 1D and Figure 2). We then hypothesized that it could be possible for the mutated receptor to still form complexes with the IL23R or IL-12Rβ2 but not bind IL-12 or IL-23 in a conformation that activates downstream signaling. The patient’s RCRS history led us to focus on IL-12/Th1 rather than IL-23/Th17 defects. To assess the potential role of the *IL12R*β*1* mutation on IL-12 signaling, we measured IFNγ production by stimulating PBMC from the patient and healthy age matched controls after IL12Rβ1 specific stimulation. The expected result for a heterozygous patient would be a reduction rather than abolishing IL-12 signaling. The results show that the percentages of the CD4^+^ T cells that are IFNγ^+^ after stimulation are 4.6% for the healthy control versus 5.2% for the patient. However, only 0.5% of the patient’s CD4^+^ T-cells are IFNγ^high^ compared to 2.8% in the healthy control (Figure 3).

**Figure 2.**
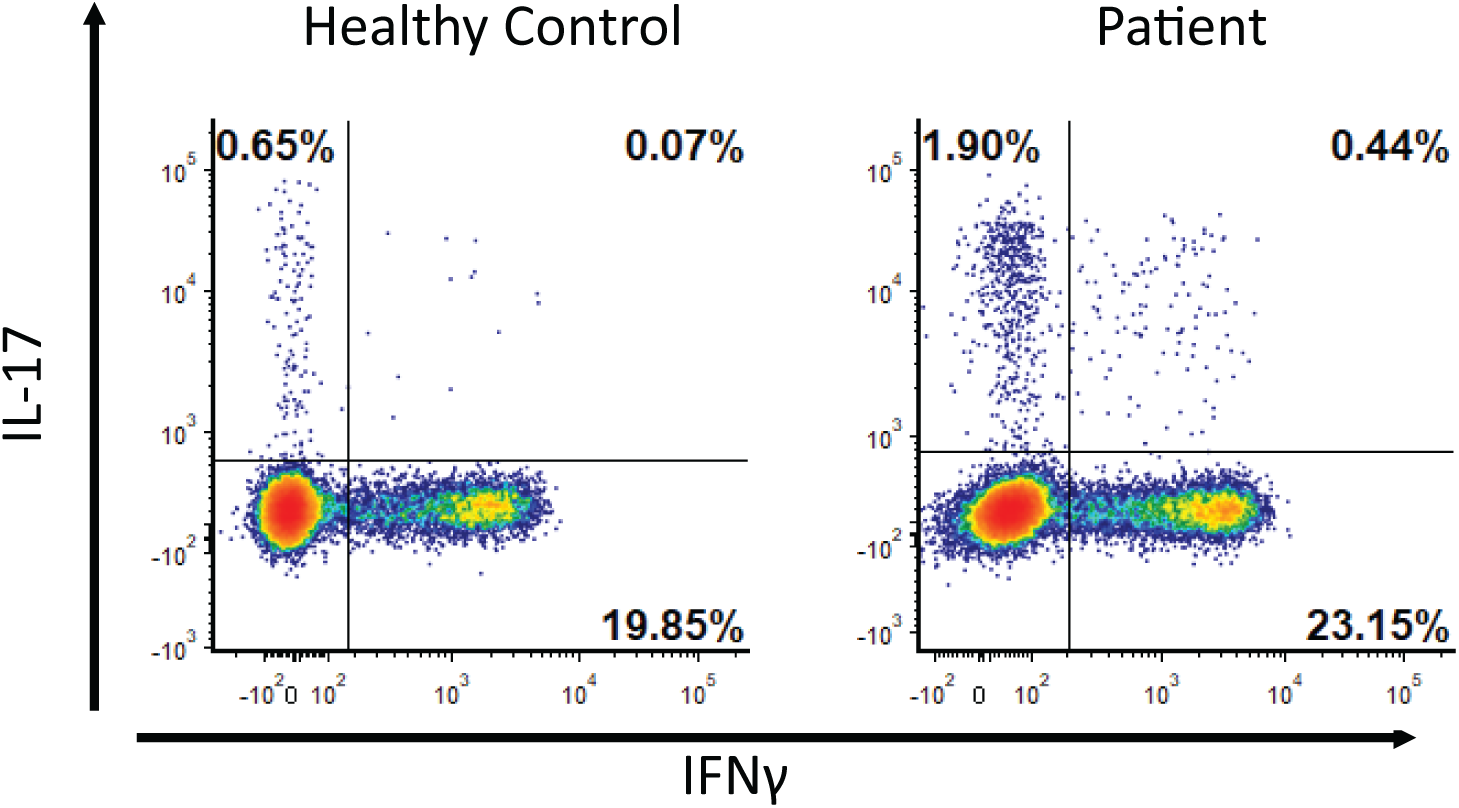
Stimulation of PBMC with PMA and ionomycin. The percentages of CD3^+^/CD4^+^ lymphocytes that produces IL-17 (normal range 0.2%-2.2%) and IFNγ (normal range 3-30%) were determined by flow cytometry.

**Figure 3.**
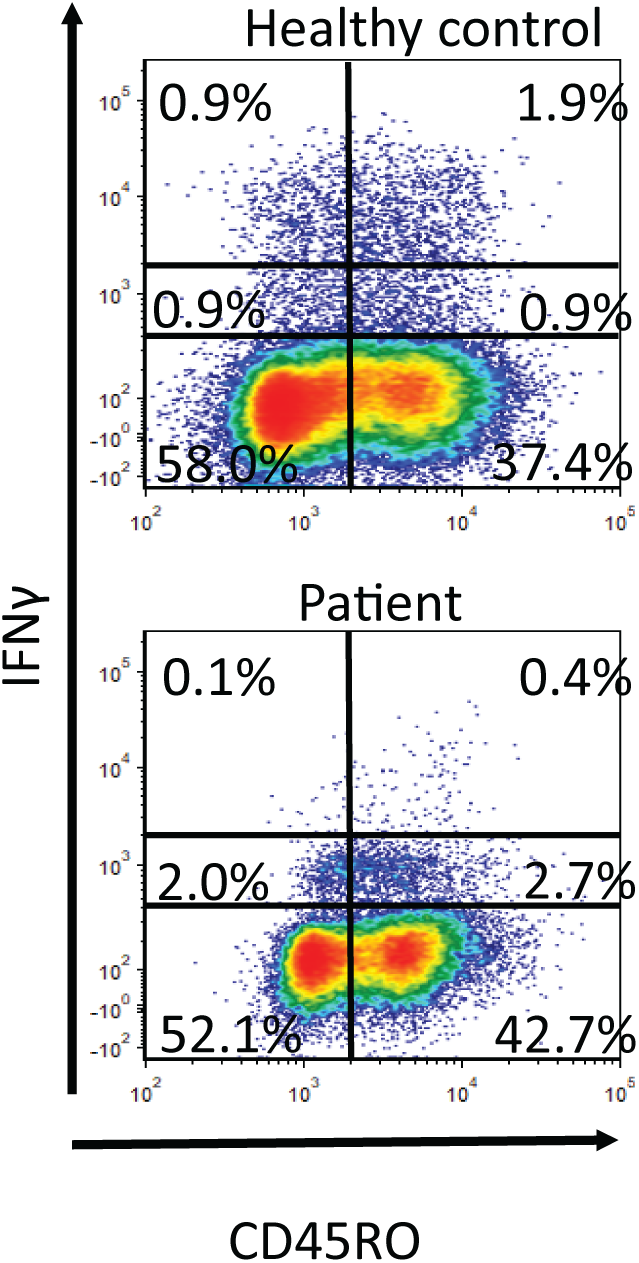
The mutation affects IL-12 signaling. PBMC stimulated with IL-12, IL-18, IL-2 and CD3&CD28 beads. The IFNγ production were measured in CD3^+^/CD4^+^ lymphocytes. The experiment was repeated three times. Representative plots are shown.

To identify the Th1 defect, we measured the phosphorylation of STAT4 in response to IL-12 stimulation in CD3^+^/CD4^+^/CD45RO^+^ cells at various time points between 0 and 5 hours. The result for the patient is an average of three samples each collected a month apart. They were compared to a group of age matched healthy controls (n=5) and patients with various well defined immune deficiencies requiring antibody replacement therapy (n=5). Healthy controls showed an increase in percentage of p-STAT4 positive T-helper memory cells after 30 minutes of stimulation with IL-12 which peaked after 60-120 minutes followed by a slow decrease after 180 minutes. The patient’s p-STAT4 response was significantly blunted at all time points. (Figure 4A and 4B).

**Figure 4.**
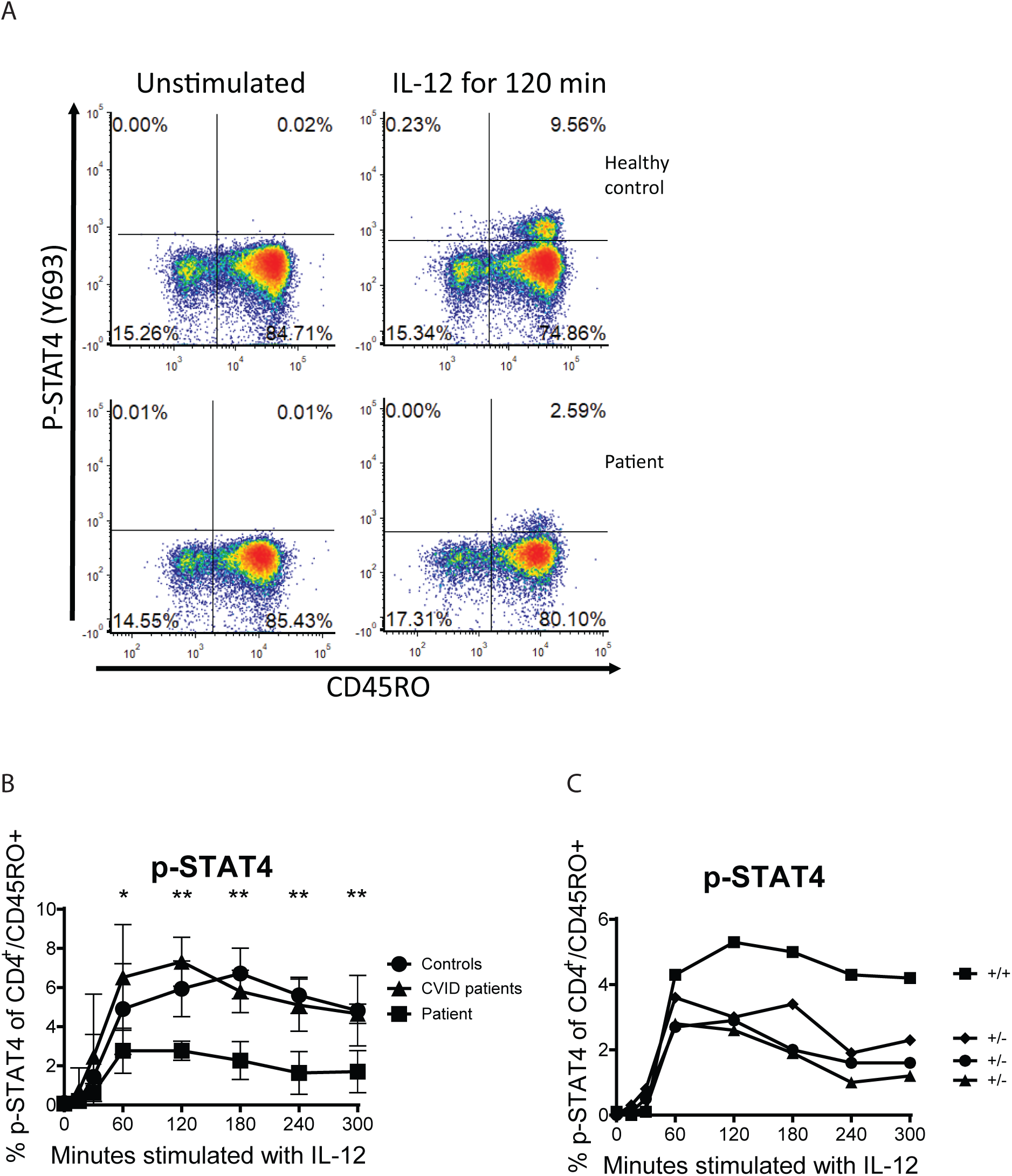
STAT4 phosphorylation in response to IL-12 stimulation. A) representative plots. B) Results from the patient, healthy controls and CVID patients. C) Four of the patient’s children. “+” indicates wt allele and “-“ indicates the mutation.

We subsequently tested four of the patient’s adult children. The IL-12 induced STAT4 phosphorylation showed that the three siblings with the mutation showed a lower peak in STAT4 phosphorylation after 60 minutes compared to the sibling without the mutation (Figure 4C). One of the three siblings with the mutation had a history of recurrent sinusitis and pneumonia.

### Transfection Experiments to show the impact of Q2883 mutation on primary T cells

The modeling of the IL12 receptor did not show any difference in the folding of the receptor between the wild type and the p.Gln238Glu variant (Supplemental Figure 1). To determine whether the mutated IL12 receptor is the cause of the defective STAT4 phosphorylation, we first engineered primary CD4^+^ T cells to overexpress the WT or the mutated IL-12Rb1 (Supplemental Figure 2). We saw no difference in the phosphorylation of STAT4 on IL12 stimulation in these primary CD4^+^ T cells when compared those engineered to express wild-type or mutant IL12Rb1. (Supplemental Figure 3). A significant challenge posed by this system is the mosaicism of the cells harboring the mutant receptors, as they also express normal IL12b1 receptors constitutively. Consequently, IL12 may exhibit a preferential activation of the normal receptor, complicating the functional analysis of the mutant variant. To address this issue, we utilized Jurkat cells, which are deficient in IL12b1. Jurkat cells also express high levels of STAT4 constitutively (Figure 5). Thus, we genetically expressed either the wild-type (WT) or the mutant variant of this receptor in Jurkat cells along with the IL-12Rb2, which is required to form heterodimer with IL-12Rb1 for the signaling via IL-12 (Supplemental Figure 4). In these genetically engineered Jurkat cells, upon stimulation with IL-12 we could detect phosphorylation of STAT4, but not wild type Jurkat line, suggesting efficient reconstitution of the receptor complex for this signaling pathway. Overexpression of IL12Rb1 and IL12Rb2 genes also led to high level expression of these receptors on cell surface of the Jurkat cells and was comparable between the mutant and the WT IL-12Rb1 expressing cells (Figure 6A). Upon stimulation of with IL-12, there was no difference for STAT4 phosphorylation between the Jurkat cells expressing WT or the mutant IL12Rb1 (Figure 6B). As we couldn’t establish a conclusive link between the patient’s mutation and the signaling defect, we opted to examine a larger number of individual clones.

**Figure 5.**
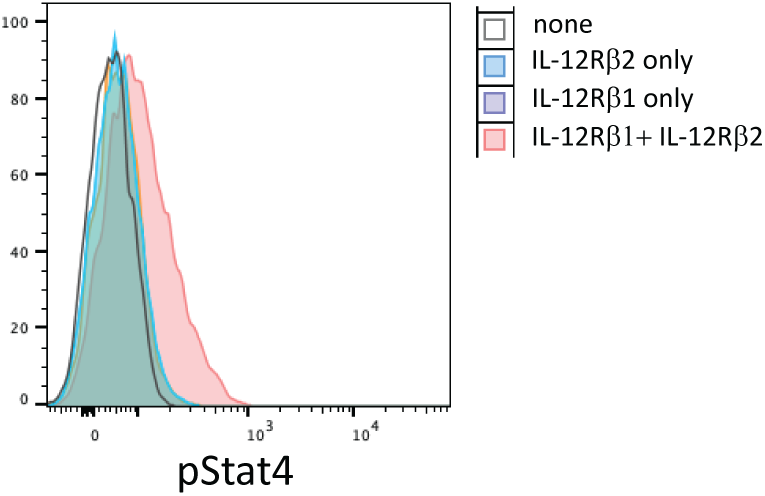
Effect of IL12Rβ1 expression on IL-12 induced STAT4 phosphorylation in Jurkat cells. Histograms overlay representing pStat4 after stimulation of wild type or engineered Jurkat cells with wt IL12Rb2, wt IL12Rb1 or both receptors.

**Figure 6.**
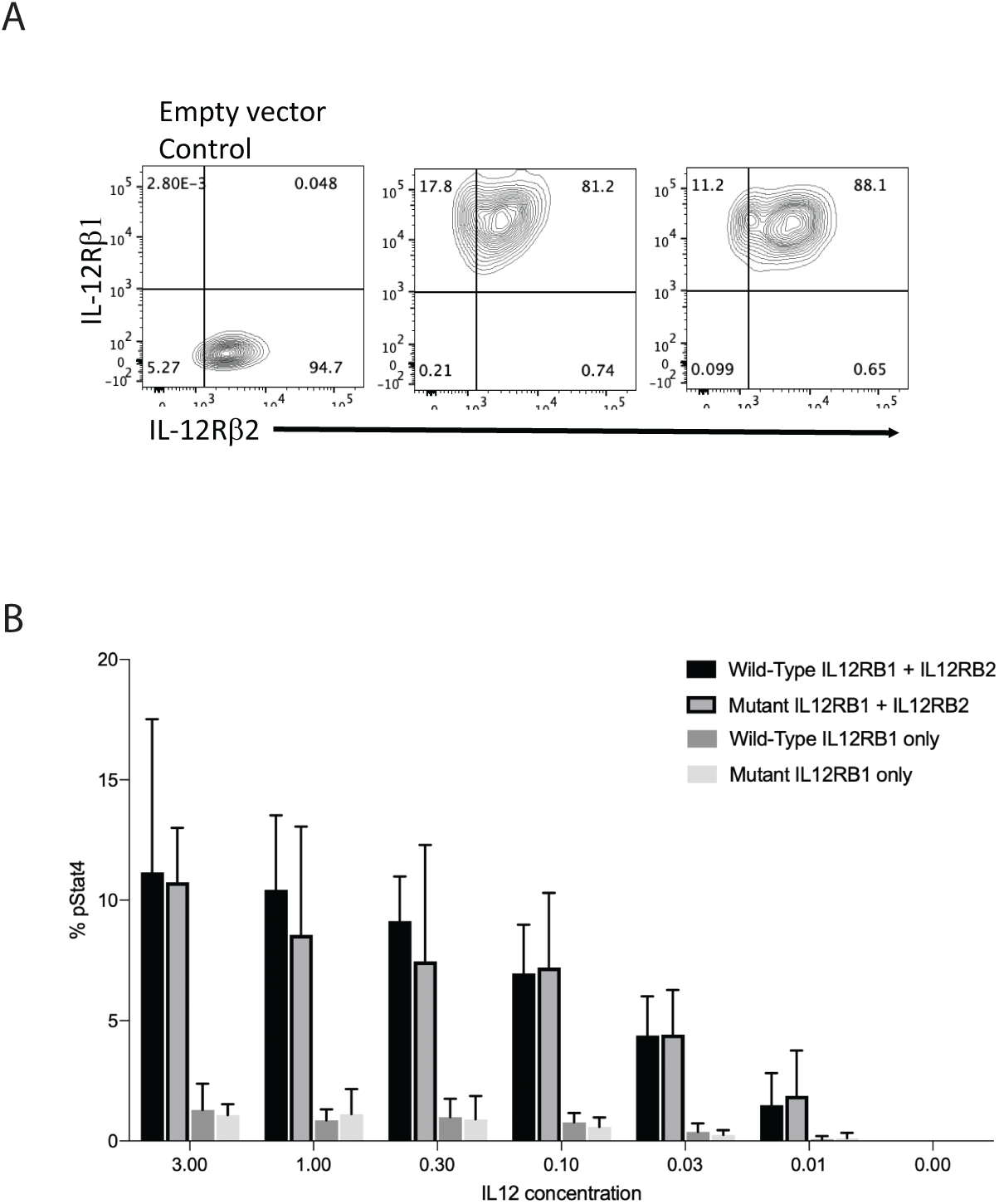
IL12Rb1 and IL12Rb2 engineered Jurkat cells. A) Expression analyzed by flow cytometry of IL12Rb2 and IL12Rb1. B) Comparison of percent phosphorylated STAT4 expression in engineered Jurkat cells expressing wild-type or mutant IL12Rb1 with or without IL12Rb2.

We therefore asked if there factors other than IL-12R and STAT4 expression could potentially affect the IL-12 signaling. We reasoned such differences could only be detected at single cell clone levels. Accordingly, Jurkat cells engineered to overexpress WT IL-12Rb2 and WT IL-12Rb1 were single cell sorted based on GFP and RFP expression, single cell clones were expanded for 3-4 weeks and pStat4 levels in different clones were measured after IL-12 stimulation. Remarkably, while the IL-12 receptor levels were high and comparable among all clones (over 40 clones were analyzed), pSTAT4 levels were widely different between clones stimulated with IL-12 (Figure 7A). These results suggest that yet to be determined intermediary factors between the receptor and STAT phosphorylation, for example kinases such as Tyk2 or Jak2, may influence the efficiency, which could explain the results observed in the family reported in this article (Figure 7B).

**Figure 7.**
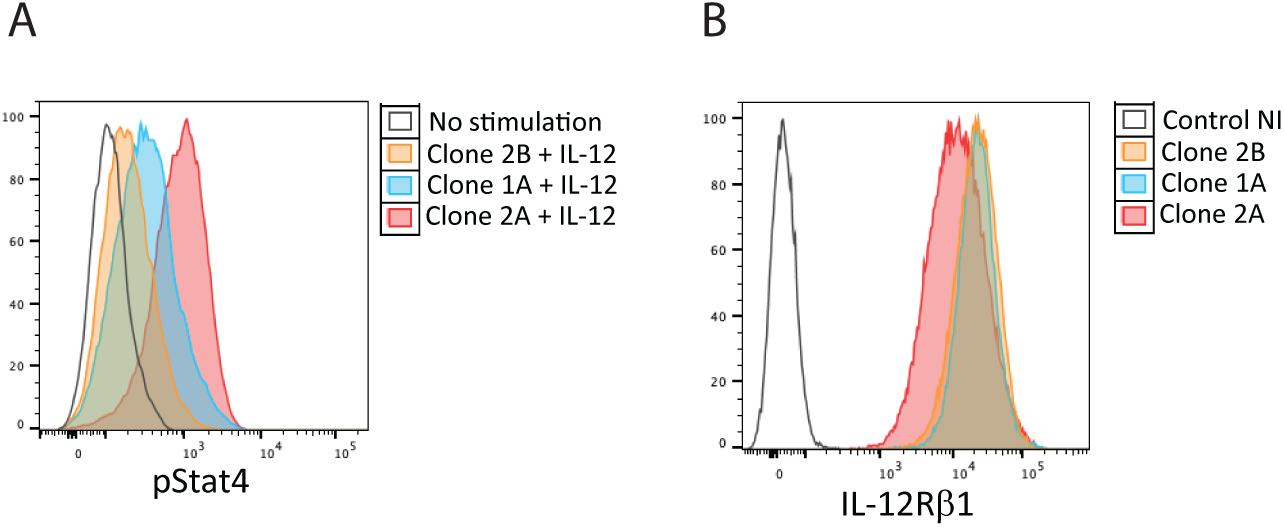
STAT4 phosphorylation in Jurkat clones expressing different levels of IL-12Rb1. A) STAT4 phosphorylation after IL-12 stimulation. B) IL12Rb1 expression levels.

### Validation of the pSTAT4 assay

In the context of CRS, dysbiosis in the upper airway microbiome can influence IL12 signaling. For example, mucin-fermenting bacteria in CRS may degrade and ferment mucins, altering the airway environment and promoting inflammation that could disrupt IL12 signaling [33]. Additionally, elevated short chain fatty acid levels in CRS mucus may also impact this pathway [34]. This suggests that apart from genetic factors, extrinsic factors, such as microbial imbalances could contribute to IL12 signaling defects in CRS.

To test this hypothesis, we next sought to test a larger cohort of patients with RCRS, to see if the variability we see in the pSTAT4 levels in the clones is reflected between patients. The phosphorylation of STAT4 after IL-12 stimulation allows for a functional readout of the signaling regardless of the location of the genetic variations in the IL-12/IFNγ axis or molecules that influence it, which can vary between patients. To do this, we first validated the assay so that the results are consistent and that it can be included in the clinical workup of patients with CRS. The previously published pSTAT4 assay, used to study IL-12 receptor mutations, relied on the hypothetical approach of maximizing IL-12 receptor expression by inducing blastogenesis using PHA, a process that typically takes 5-7 days. The blasts are then subsequently stimulated with IL-12 [35–37]. This approach can be employed in a research setting to demonstrate that IL-12 receptor mutations can disrupt IL-12 signaling. However, it is not well-suited for routine clinical testing due to the challenges associated with validating a multistep process that involves numerous variables in blastogenesis. Additionally, blasts may not accurately reflect a physiological response in vivo, as the majority of circulating cells are not in a continuous activated state. Furthermore, our findings indicate that the expression of T cell surface IL-12 receptors remains unchanged between resting T cells and blasts. (Supplemental Figure 5). The pSTAT4 assay performed on resting primary T cells and validated under CAP/CLIA utilizes whole blood within 24 hours [26]. The assay measures STAT4 phosphorylation at multiple time points, extending up to 2 hours, and can be finalized within 5 hours. It exhibits high reproducibility and is accompanied by reference ranges, enabling us to assess a patient’s condition without the need to include a group of healthy controls.

### pSTAT4 to screen CRS for prolonged surgical recovery

In addition to the results from our Jurkat clones showing variability in STAT4 phosphorylation, the published reports on T cell defects in up to 56% of RCRS patients supported our approach to screen a larger cohort of RCRS patients [38]. To assess the clinical relevance of our finding in the larger RCRS population, we used the IL-12 induced pSTAT4 as a screening assay in 147 RCRS patients requiring surgery (Figure 8 and Supplemental Figure 6). These RCRS patients presented with infections of Klebsiella, Staphylococcus, Serratia and Burkholderia species isolated from sinus cultures, and had history of multiple sinus procedures, no polyps. These patient’s nasal mucosa showed areas of inflammation, often with one side or another showing much worse disease, rather than widespread inflammation. Chronic inflammatory infiltrates were common to all RCRS patients’ tissue specimens, performed as a part of their post-surgical specimen processing. Bad surgical outcome was defined as requiring more than two courses of antibiotic therapy within the first 6 months after surgery. Low STAT4 phosphorylation correlated with prolonged surgical recovery of lengthy healing process as well as persistence of the sinus infections. None of the individuals in the control group or the patients with a good surgical outcome had a pSTAT4 value below 5% whereas most of the patients with poor surgical outcome had a value below 5%. To test if a soluble factor (i.e. anti-cytokine antibody) was responsible for low STAT4 phosphorylation, we performed the experiments on health subjects CD4 T cells using RCRS patient serum. Serum of RCRS patients with low pSTAT4 did not result in healthy patients’ cells down-regulating STAT4 phosphorylation in response to IL-12 stimulation (Supplemental figure 7).

**Figure 8.**
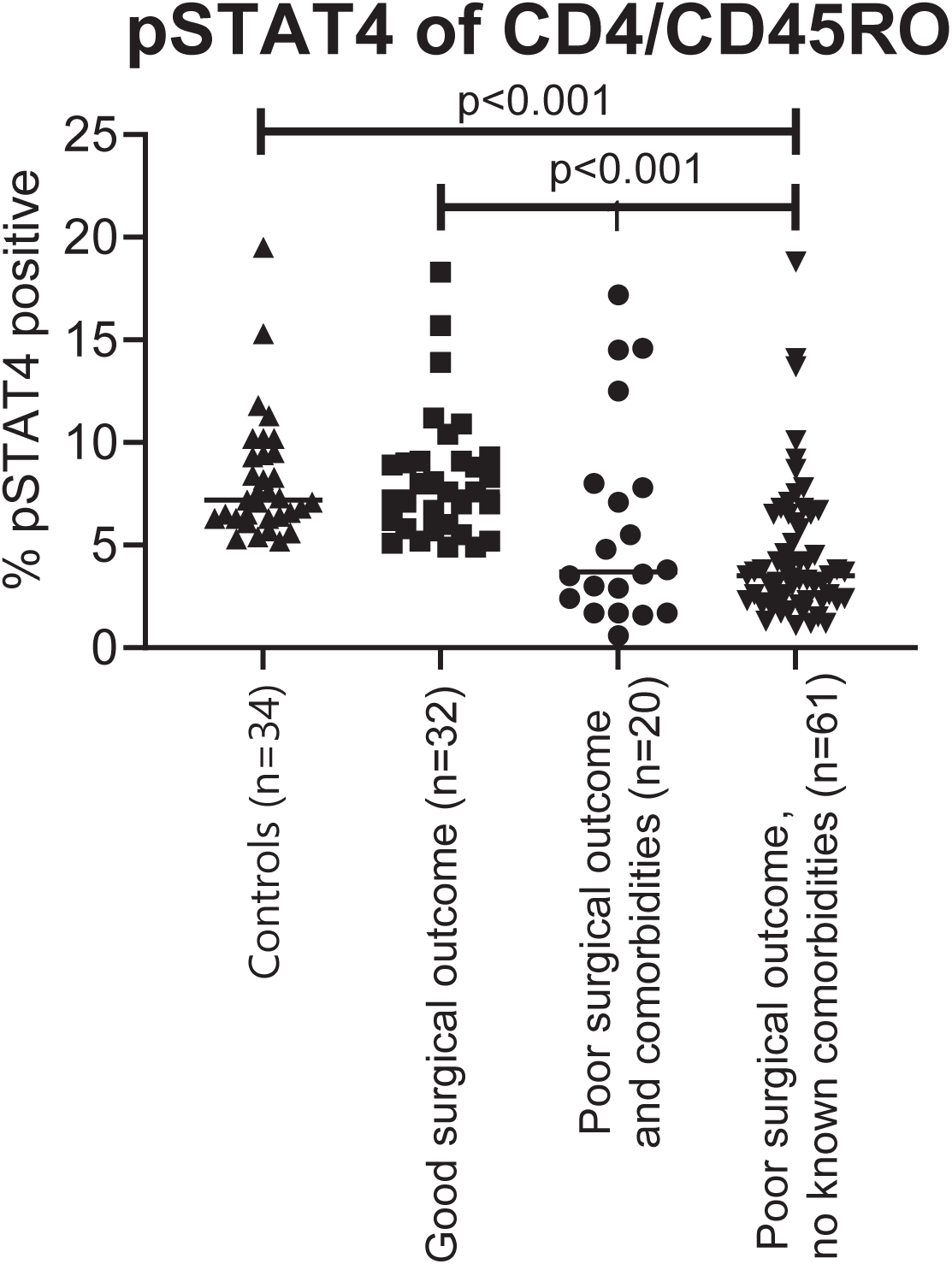
Percent pSTAT4 positive cells after IL-12 stimulation of whole blood from CRS patients and healthy controls.

### Effect of Lactobacilli on STAT4 phosphorylation in-vitro and in vivo

In our follow-up experiments, we explored the potential impact of lactobacilli on pSTAT4 phosphorylation. This investigation was prompted by data suggesting that specific lactobacilli strains can trigger IL-12 production in macrophages and by anecdotal observations within our group regarding the potential clinical advantages of probiotics in chronic sinus disease. Our results demonstrated that when PBMCs were exposed to a lactobacilli solution containing L. acidophilus for 18 hours, STAT4 phosphorylation was induced, even in the absence of exogenous IL-12 supplementation. Intriguingly, when the lactobacilli solution was introduced to PBMCs of patients exhibiting low STAT4 phosphorylation in their CD4 T-cells, there was a notable enhancement in STAT4 phosphorylation. This effect was partially inhibited by neutralizing anti-IL12 antibodies (Figure 9). This suggests that the probiotic’s effect can be attributed to IL12 production but contributions from an IL-12 independent pathway could not be ruled out. The phosphorylation of STAT4 by lactobacilli, as well as its inhibition of IL-12, exhibits specificity to both the bacterial strain and the host (Supplemental Figure 8).

**Figure 9.**
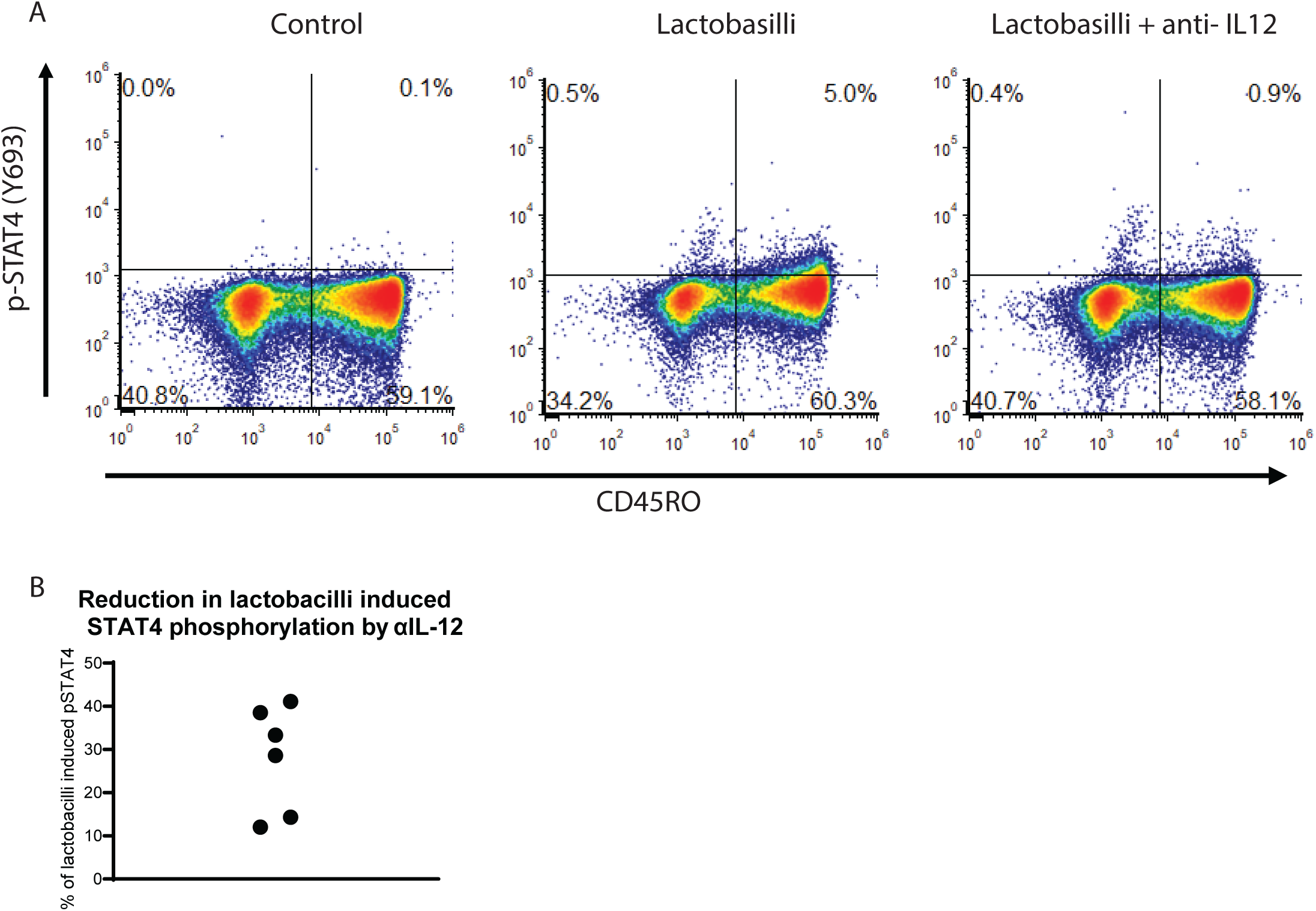
*In vitro* effects of lactobacilli on STAT4 phosphorylation. A) The percentage of pSTAT4/CD45RO cells for the different conditions indicated in the figure. B) Effect of IL-12 neutralizing antibodies on lactobacilli induced STAT4 phosphorylated.

In light of these findings, we conducted a retrospective review of medical records for 25 CRS patients who were administered *Latilactobacillus sakei*, a strain reported for its ability to induce exceptionally high IL-12 levels [39]. We gauged the patients’ response to the probiotic by correlating the number of postoperative office visits and change in pSTAT4 levels pre-and post-probiotic use in a 1-year period. The results present pSTAT4 levels as a percentage change, and the outpatient data reflect the difference in visit frequency. Our analysis underscored a positive correlation between the frequency of patients’ post-surgical visits and the pSTAT4 phosphorylation in their CD4 T cells (Figure 10).

**Figure 10.**
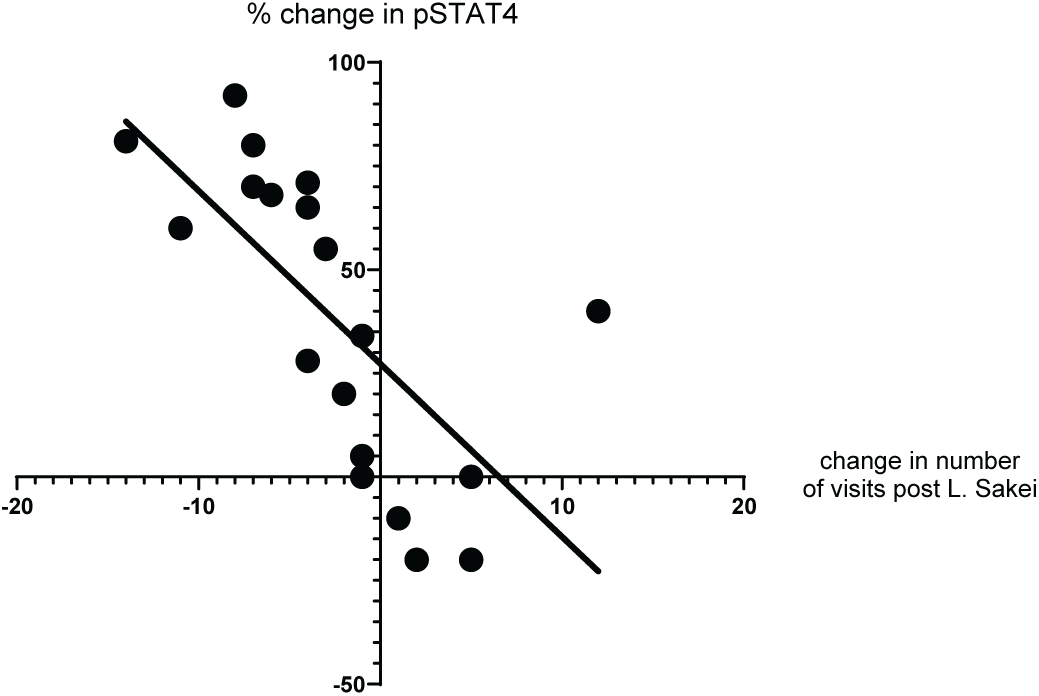
Lactobacilli in post-surgery setting. Patient data on the total number of outpatient visits for one year following surgery and pSTAT4 levels before and approximately six months after *L. sakei* treatments.

## Discussion

CRS remains a nuanced pathology, both in its presentation and underlying etiologies. In this study, we started out with exploring a VUS in the IL-12 receptor beta 1 of an RCRS patient. Despite the patient’s receptor expressing normally on peripheral blood T-helper cells, our discoveries highlighted a compromised IL-12 signaling. Even though there is a correlation in the patient’s family between individuals with the mutation and low pSTAT4 response we were unable to show an effect of the mutation in computational structural simulation models or by *in vitro* transfection experiments.

Broadening our scope, we undertook a validated pSTAT4 assay across an expansive cohort of RCRS patients. The outcome was illuminating; diminished STAT4 phosphorylation in CD4 T-cells heralded poor post-surgical recoveries. While this finding seemed to be a validation of prior research emphasizing T-cells’ pivotal roles in RCRS, it also accentuated a conspicuous gap in clinical practice: the lack of routine cellular immunity assessments of these patients, despite their potential diagnostic importance. The common practice of immune evaluation of CRS involves measuring serum quantitative antibody levels, which, more often than not, provide inconclusive results [25,40,41]. As this research underscores specific defects in T cell function and offers a clinically validated diagnostic tool, we anticipate a paradigm shift towards more tailored therapeutic approaches for RCRS. However, unveiling the exact mechanics behind impaired IL-12-mediated STAT4 phosphorylation remains to be determined, the deciphering of which could usher in therapeutic breakthroughs.

Another riveting aspect of our study revolves around the interplay between Lactobacillus species and the immune system. With their presence predominantly in the human gut, Lactobacillus species’ interactions with macrophages can influence cytokine profiles, including IL-12 [42]. We confirmed this in our *in vitro* analysis using neutralizing IL-12 antibodies which reduced the effect of *L. acidophilus*. Furthermore, the *L. acidophilus* was able to induce STAT4 phosphorylation in the absence of any exogenous IL-12. But not all Lactobacilli are created equal. Potentially, strain-specific variability can exist in their potency to modulate IL-12, potentially attributed to differences in their cell walls [43]. This study further delves into this variability, reinforcing the importance of strategic Lactobacillus selection in potential therapeutic endeavors. The precise mechanism of Lactobacillus stimulation of T cells is not entirely known and our data shows both IL-12 independent as well as dependent phosphorylation of STAT4.

The STAT4 phosphorylation we observe in Figure 9A is lower than what we see when saturating the blood sample with IL-12, but it is likely to be adequate, *in vivo*, to activate and poise the Th-1 cells in patients with low pSTAT4 response and lead to improved clinical outcomes in postsurgical RCRS patients.

Our Lactobacillus data provide important insights into the inconsistent findings in the literature concerning the clinical effects of Lactobacillus on Chronic Rhinosinusitis (CRS) outcomes. The data highlight two key factors: Firstly, the specific strains of probiotics and individual host characteristics significantly influence STAT4 phosphorylation. This effect’s strength and its IL-12 dependence vary, as evidenced by the inhibition assays presented in Figure 5 and Supplementary Figure 8. Future research should identify the probiotic structural components responsible for these effects. Secondly, the choice of clinical endpoints is crucial. Given that STAT4 phosphorylation is pivotal in regulating Th1 cell responses, our research prioritized post-surgical wound healing rather than improvements in clinical symptoms or reductions in sinus infection rates. [44–46]. The process of wound healing begins with an immediate inflammatory response, seeing an influx of various cellular actors. Th1 cells, through their influence on macrophages and fibroblasts and their production of IFN-γ, impact collagen synthesis, the bedrock of tissue repair [47]. However, maintaining a balance in Th1 activity is paramount. An overwhelming or inadequate Th1 response can either prolong inflammation or inadequately support the initial healing phases, respectively. As such, understanding Th1 activity, primarily via STAT4 phosphorylation, provides invaluable insights into predicting and optimizing surgical outcomes. Furthermore, the microbiota can induce an adaptive immune response that can couple antimicrobial function with tissue repair and wound healing [48]. It has been shown that non-classical MHC class I molecules, an evolutionarily ancient arm of the immune system, can promote immunity to the microbiota, such as Lactobacillus, and subsequently affect cytokine signaling, including IL-12 [49]. To enhance our understanding of Recurrent Chronic Rhinosinusitis (RCRS), it is crucial to conduct prospective studies focusing on the IL-12 pathway. Investigating its specific functions could provide insights into its dual role in infection control and inflammation regulation.

A limitation of our study is that it was not designed prospectively, and it depended on the analysis of retrospective review of patient charts. To further corroborate our clinical findings, future studies should be prospective, randomized, and placebo controlled.

Finally, our research underscores the critical role of Laboratory Developed Tests (LDTs) in enhancing clinical diagnostics and patient care, with a special focus on the pSTAT4 assay as an example. Unlike test kits cleared by the Food and Drug Administration (FDA), LDTs bridge the gap in diagnosing complex or rare conditions. Their ability to rapidly integrate cutting-edge diagnostic methods makes them an essential tool in the dynamic healthcare environment, especially when leveraged by skilled clinicians. The pSTAT4 test, in particular, has shown promise as a predictor of outcomes following sinus surgeries, potentially revolutionizing the management strategies for RCRS. Given these findings, there is a compelling case for the broader adoption of LDTs like pSTAT4 in clinical settings and a call for more research to further establish their utility and impact on patient care trajectories.

## Conclusion

As our grasp of RCRS and its complex underlying mechanisms grows, evaluating T-cell defects using the STAT4 phosphorylation assay described in this study emerges as a particularly promising avenue. These innovative diagnostic and therapeutic strategies that evolve from them, have the potential to transform our management of CRS. When utilized with precision, they offer a path toward alleviating the substantial strain that CRS places on healthcare systems.

## Data Availability

All data produced in the present study are available upon reasonable request to the authors

