## Supplementary material for "STAT4 Phosphorylation of T-helper Cells predicts surgical outcomes in Refractory Chronic Rhinosinusitis": suppl figures

**Supplemental figure 1**

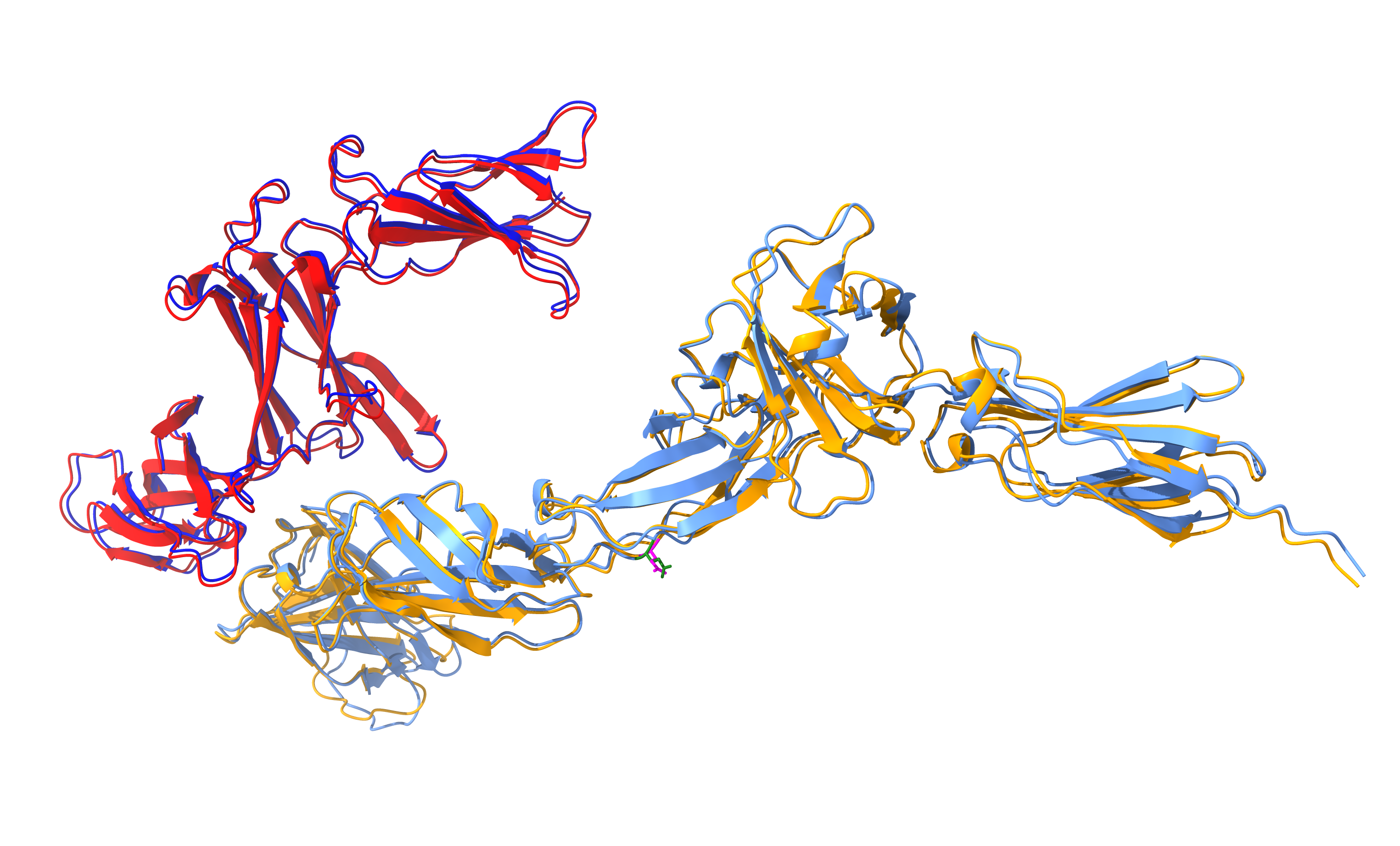

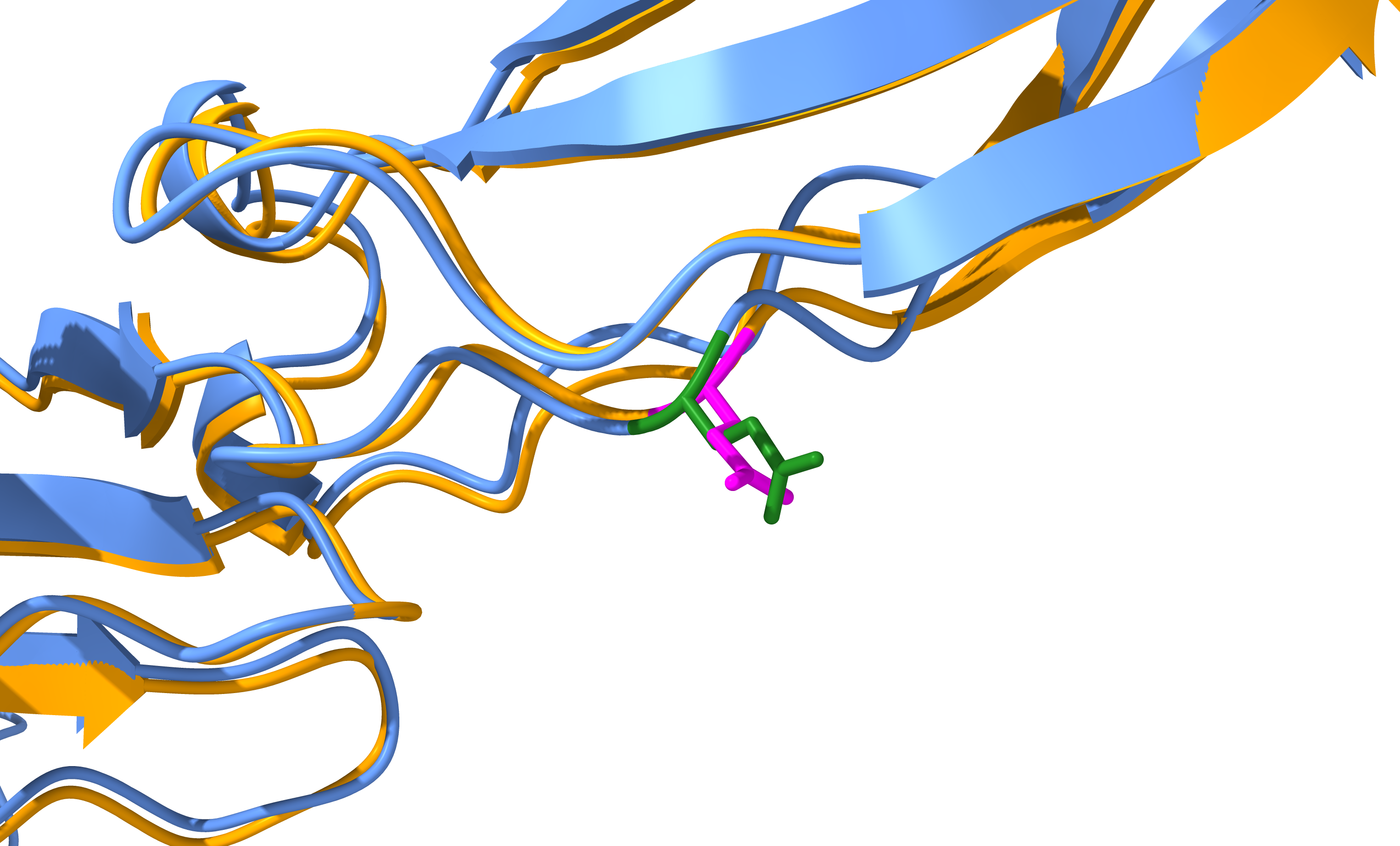

w.t.

p.Gln238Glu

IL-12β

IL-12Rβ1

aa238

IL-12Rβ1 residue 238 with the p.Gln238Glu variant was located in the linker region between two domains and distant from the interface between IL-12β and IL-12Rβ1.

IL-12β and both the wild-type and the p.Gln238Glu variant of IL-12Rβ1 were modeled using AlphaFold, and the resulting structures were aligned and visualized using ChimeraX. Residue 238 of IL-12Rβ1 was distant from the protein-protein interface and was pointed in the opposite direction. This single amino acid polymorphism did not globally perturb the protein complex structure and cased on this computer model it is likely that the Q238E variant affects the protein-protein interaction indirectly.

**Supplemental figure 2**

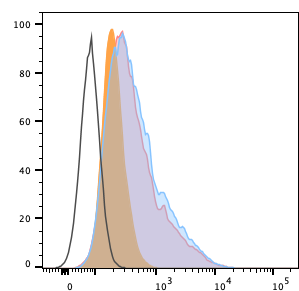

IL-12Rβ1

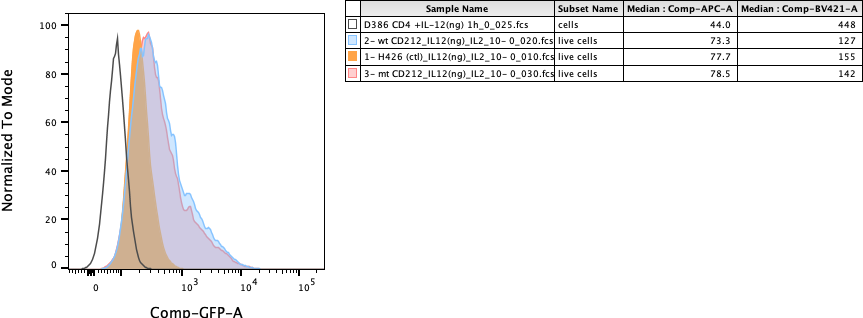

wt IL-12Rβ1

mt IL-12Rβ1

unstained

wt and mutant IL-12Rb1 overexpressed in Primary CD4 T cell

IL-12Rb1 expression in engineered primary CD4 T cells. Histogram overlay of IL-12Rb1 expression in CD4 T cells transduced with Empty vector control, wt IL-12Rb1 or mt IL-12Rb1, analyzed by flow cytometry. Empty histogram represents Fluorescence background in unstained cells.

**Supplemental figure 3**

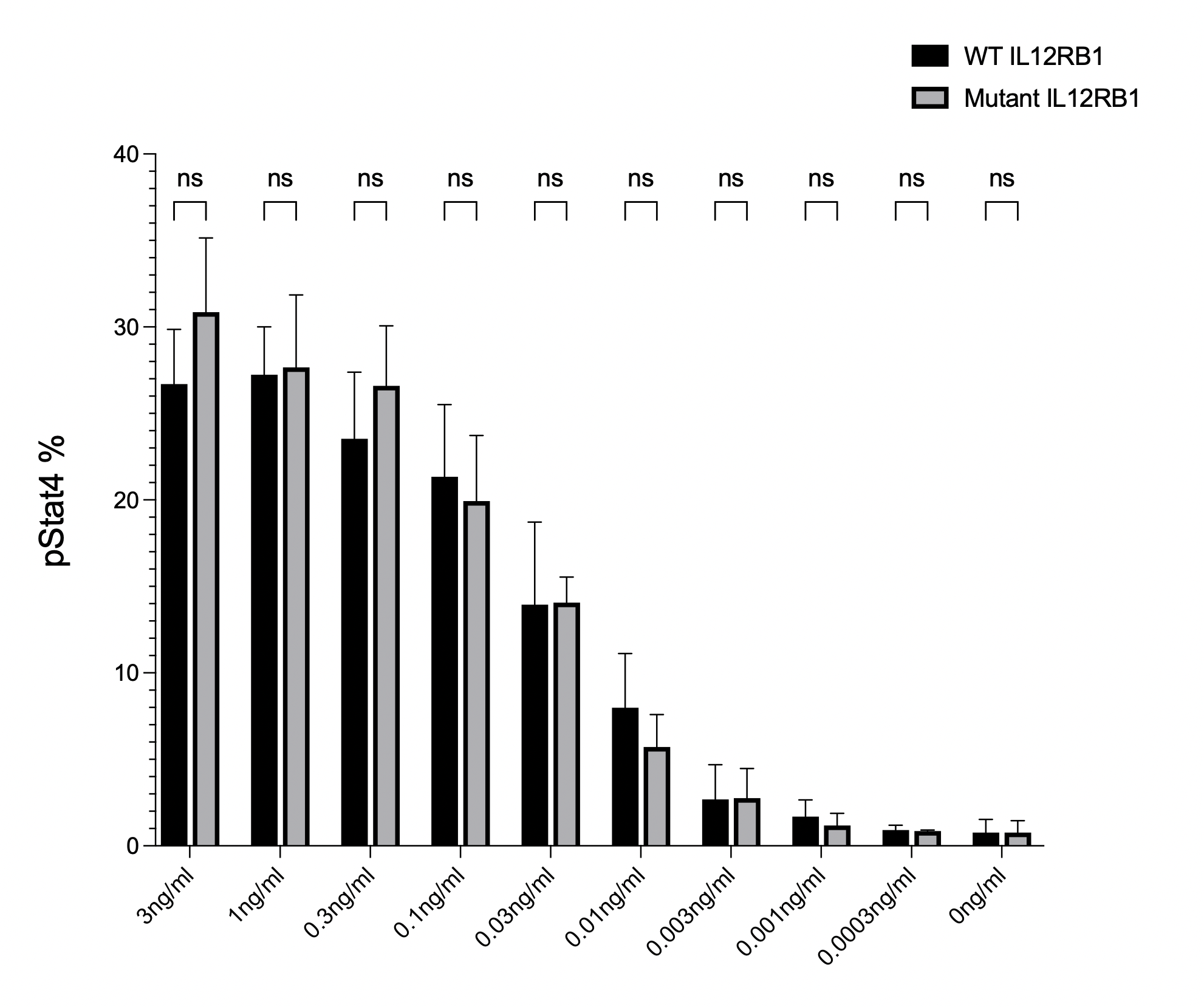

IL-12

Comparison of percent phosphorylated STAT4 expression in primary CD4 T cells engineered to express wild-type or mutant IL12Rb1. Bar graph of Phosphorylated Stat4 (pSTAT4) expression in CD4 T cells transduced with wild-type IL-12Rb1 or mutant IL-12Rb1, stimulated with different concentrations of IL12, and analyzed by flow cytometry. Non-parametric t-tests were used to determine the statistical significance. No statistical significance was observed.

**Supplemental figure 4**

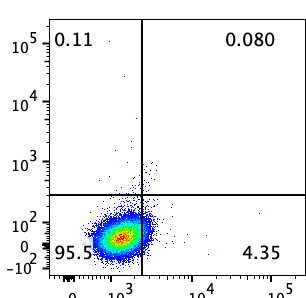

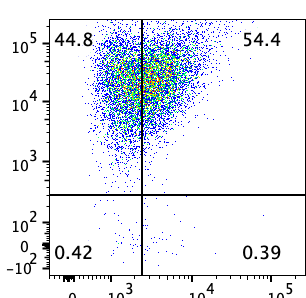

IL-12Rβ2

IL-12Rβ1

Non transduced

wt IL-12Rβ2+

wt IL-12Rβ1

IL-12Rb1 and IL-12Rb2 expression in engineered Jurkat cells. Jurkat cells non transduced or transduced with wt IL-12Rb2 plus wt IL-12Rb1 were analyzed by flow cytometry for the expression of IL-12 receptors IL-12Rb2 and IL-12Rb1.

**Supplemental figure 5**

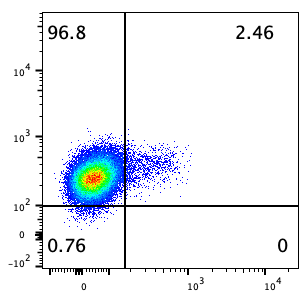

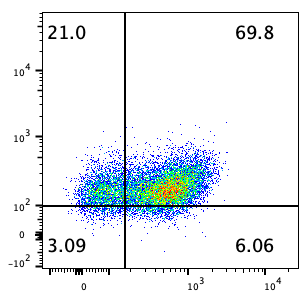

resting

activated

IL-12Rβ1

pStat4

IL-12Rb1 expression and IL-12 induced Stat4 phosphorylation in resting and activated CD4 T cells. Flow cytometry analysis of IL-12Rb1 expression and Stat4 phosphorylation in healthy donors’ PBMCs resting or day 14 post activation after stimulation with 1ng/ml IL-12. Cells were gated on live CD3+CD4+ population. Data is representative of 2 different donors.

**Supplemental figure 6**

| Characteristic | Control group | Good surgical outcome | persistence of recurrent infections | |
| --- | --- | --- | --- | --- |
|  |  |  | No known comorbidity | known comorbidities |
| Age | 34 (11-80) | 50 (21-90)^*^ | 54 (15-77) ^*^ | 55 (24-70) ^*^ |
| Sex (male/female) | 17M/17F | 12M/20F | 23M/38F | 9M/11F |
| %pSTAT4 of CD4^+^/CD45RO^+^ | 8.2 (5.2-19.5) | 8.2 (4.9-18.3) | 4.5 (1.1-18.8) ^**,#^^#^ | 5.9 (0.6-17.2) |
| IgG (mg/dL) | 997 (405-1595) | 1088 (746-2154) | 1007 (525-1492) | 1054 (391-1634) |
| IgA (mg/dL) | 180 (5-606) | 207 (74-357) | 228 (47-700) | 177 (45-498) |
| IgM (mg/dL) | 71 (5-159) | 107 (32-312) ^*^ | 101 (28-333) ^*^ | 104 (22-275) |
| %CD3^+^ | 81 (42-97) | 77 (62-87) | 75 (48-89) | 71 (19-88) |
| #CD3^+^ | 1458 (702-2798) | 1504 (502-2584) | 1410 (658-2465) | 1001 (111-1659) |
| %CD20^+^ | 12 (0-32) | 11 (2-20) | 11 (2-22) | 11 (2-27) |
| #CD20^+^ | 238 (0-606) | 216 (26-617) | 210 (21-456) | 146 (25-541) ^*,#^ |
| %CD3^-^/ CD16^+^•CD56^+^ | 13 (2-44) | 11 (4-22) | 12 (4-37) | 18 (7-61) ^#^ |
| #CD3^-^/ CD16^+^•CD56^+^ | 239 (29-1061) | 191 (67-398) | 213 (31-776) | 221 (84-451) |
| %CD4^+^ | 58 (25-71) | 48 (33-64) | 52 (30-65) | 46 (2-77) |
| %CD8^+^ | 21 (8-38) | 24 (9-39) | 20 (10-39) ^#^ | 20 (1-57) |
| %CD4^+^/CD45RO^+^ | 25 (11-53) | 25 (6-42) | 31 (11-60) ^*,#^ | 33 (6-59) ^*,#^ |
| %CD8^+^/CD45RO^+^ | 5 (0-17) | 6 (1-18) | 6 (1-25) | 7 (1-20) |

Legend: The patients undergoing surgery were divided into three groups depending on the outcome of the surgery. 1) Good outcome, no persistence infections. 2) Persistence of infections but no known comorbidities and 3) Persistence of infections and known comorbidities including CVID, SAD, Heavy smoker, low CD4 or CD8 count, or autoimmune disease. The absolute number of cells is pr μL. The percentage values are of all lymphocytes unless otherwise stated. * p< 0.05 compared to control group, ** p<0.001 compared to control group, # p< 0.05 compared to good surgical outcome group, ## p<0.001 compared to good surgical outcome group; students t-test.

**Supplemental figure 7**

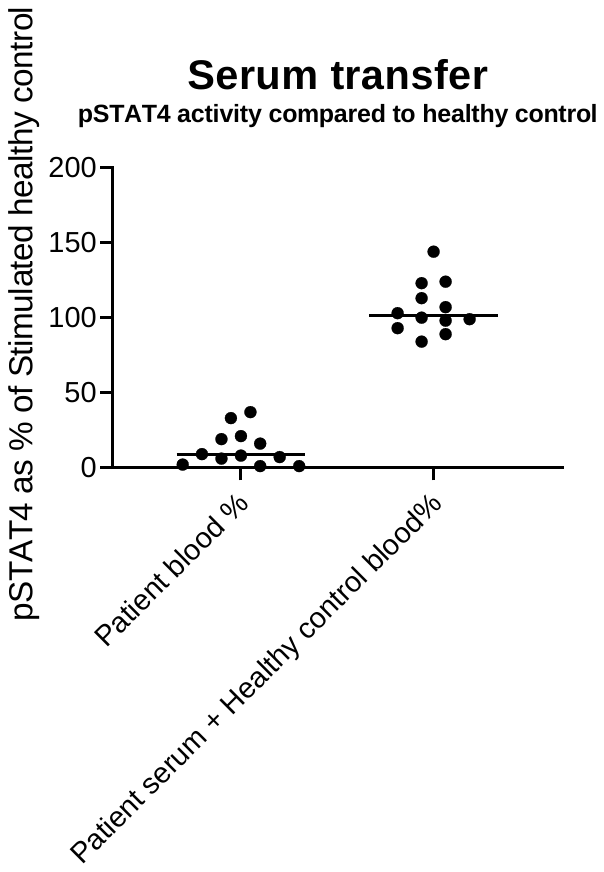

Effect of serum from a low responder on the response in a healthy control. Blood was collected from 12 patients from the group “persistence of recurring infections with no known underlying condition” All patients had a pSTAT4 response below 5%. The patients’ serum was then mixed with blood from a healthy control. The first column shows the response in 12 different patients as a percentage of the positive (healthy control). The second column shows the response in the healthy controls cells when mixed with serum from the low responding patients. The results are shown as percent pSTAT4 positive CD4/CD45RO lymphocytes after 60 min of IL-12 stimulation. All results are normalized to the healthy control.

**Supplemental figure 8**

|  | Patient 1 | Patient 2 |
| --- | --- | --- |
| Control | 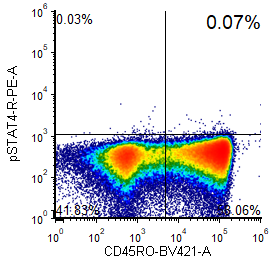 | 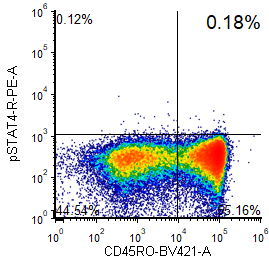 |
| Super 8 | 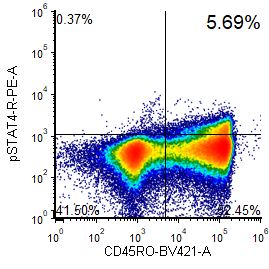 | 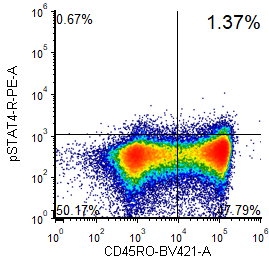 |
| Super 8 + AB | 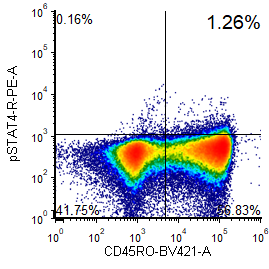 | 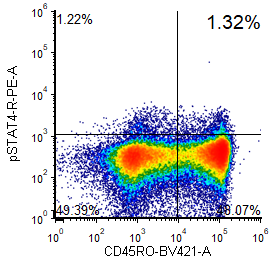 |
| Daily Care Probiotic | 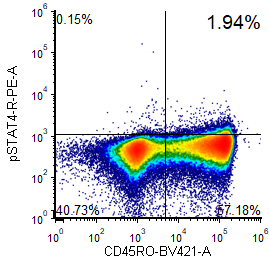 | 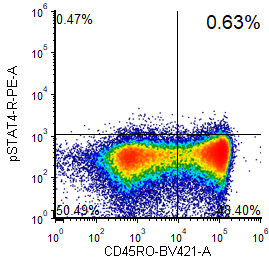 |
| Daily Care Probiotic + AB | 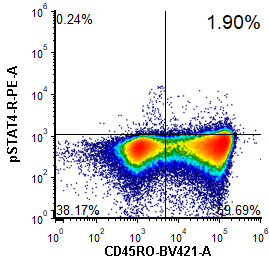 | 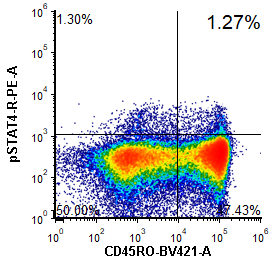 |
| Lanto Sinus | 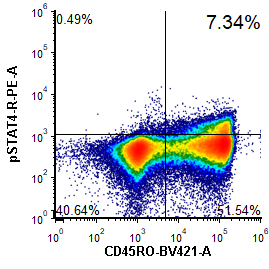 | 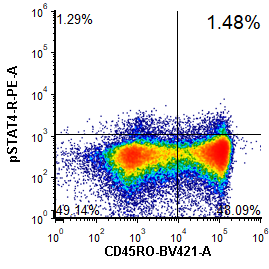 |
| Lanto Sinus + AB | 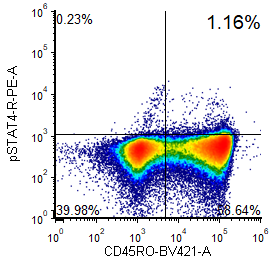 | 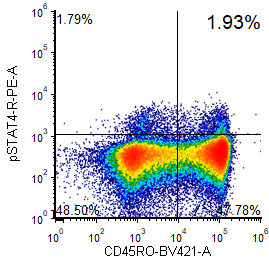 |

*In vitro* and *in vivo* effects of different Lactobacillus products. PBMC were isolated and incubated in the presents of 10^6^ CFU/ml with or without IL-12 neutralizing antibody for 18 hours at 37˚C after which the PBMC were fixed, permeabilized, stained and acquired. The initial gating was on CD3^+^/CD4^+^ lymphocyte singlets. The plots show the percentage of pSTAT4/CD45RO cells after initial CD3/CD4 gating for the different conditions indicated in the figure. The experiment shows results from two distinct different donors. SUPER 8 is a mix of 8 different probiotics where 6 are lactobasilli. Daily Care Probiotic is a mix of 6 different Bifidobacterium and Lanto Sinus only contains Lactobasillus Sakei.
